# Predicting the median lifespan of ITNs using an area-measure of risk of physical durability: proof of concept of the Risk Index

**DOI:** 10.64898/2026.05.21.26353786

**Authors:** Stephen Poyer, Julie-Anne Akiko Tangena, Frank Mechan, Matt Worges, Eleanore Sternberg, Hannah Koenker, Christen Fornadel, Albert Kilian

## Abstract

**Background:** The lifespan of insecticide-treated nets (ITNs) varies widely across settings, reflecting both intrinsic product characteristics and external factors related to use, care, and environment. While the resistance to damage (RD) score captures intrinsic product durability, there is no standardized metric to quantify contextual risks. This study presents a proof of concept for the Risk Index (RI), a composite measure of site-level risk factors for ITN physical durability and survival.

**Methods:** We conducted a secondary analysis of durability monitoring data from 44 sites across 15 countries in sub-Saharan Africa, covering 14 ITN products. The RI was calculated as a weighted composite of 12 indicators spanning net handling, net care attitudes, and use environment. Associations between RI and median ITN survival were assessed using weighted linear regression and multivariable mixed-effects models adjusting for RD score, with country included as a random effect.

**Results:** RI scores ranged from 25.1 to 83.7 across study sites. In bivariable analysis, a 10-point decrease in RI was associated with a 4.0-month increase in median ITN survival (95% CI: 1.7–6.3; p=0.001). In multivariable analysis adjusting for RD, this association remained significant but attenuated to 2.2 months (95% CI: 0.1–4.2; p=0.037). Independently, a 10-point increase in RD score was associated with a 3.5-month increase in survival (95% CI: 1.3–5.7; p=0.001). No interaction was observed between RI and RD. Predicted survival differed by approximately one year between the lowest- and highest-risk settings.

**Conclusion:** The RI provides a standardized measure of contextual risk factors affecting ITN lifespan, independent of ITN product type. When used alongside a product’s RD score, the RI enables improved interpretation of expected site-level variation in net performance. This combined framework offers a practical basis for incorporating behavioural and environmental risk into vector control planning and for tailoring ITN strategies to local conditions.

## Background

Over the past two decades, vector control has been one of the primary drivers of global progress against malaria. Insecticide-treated nets (ITNs) accounted for 72% of malaria cases averted between 2000 and 2024, corresponding to an estimated 1.13 billion cases prevented [1]. Despite these gains, malaria remains a major public health challenge. In 2024, approximately nine million more cases were reported than in 2023, with an estimated 282 million cases and 610,000 deaths reported worldwide [2]. At the same time, donor funding for malaria control is declining [2,3], increasing the need to use available resources as efficiently as possible.

A major operational challenge with ITNs is that the length of their useful life varies greatly across subnational areas and, on average, is likely to be shorter than the three years typically used as the frequency for mass ITN distribution campaigns. Modelling by Bertozzi-Villa and colleagues on data from 2000-2020 estimated an average national ITN lifespan of 1.6 years in sub-Saharan Africa [4]. Localised ITN durability monitoring studies conducted since 2015 have shown that median survival varies from 18 months to six years, with a median of 2.7 years across 37 monitoring sites in sub-Saharan Africa [5]. Understanding why performance varies so greatly between sites and being able to predict likely performance would help optimize vector control planning and ITN deployment strategies in resource-constrained settings.

ITN performance is partly determined by a product’s physical specifications, including the snag strength, bursting strength and resistance to hole enlargement. These characteristics have been combined in the resistance to damage (RD) score, which has been shown to predict ITN lifespan, with higher RD scores associated with longer survival [6,7]. In addition to an ITN’s physical properties, empirical results suggest that lifespan is strongly influenced by the context in which nets are used. ITN durability monitoring studies show that survival outcomes can vary significantly for the same ITN product (brand) within and across countries. For example, the median survival for pyrethroid-only DawaPlus® 2.0 brand ITNs ranged from 1.6 - 5.3 years for data collected at nine sites in five countries [5]. Substantial variations in survival are also apparent between sites for PBO-pyrethroid and chlorfenapyr-pyrethroid ITN brands with sufficient data (Figure S1, supplementary information).

Standard durability monitoring surveys were developed to identify the key drivers of ITN performance in the field [8]. Factors commonly associated with ITN survival in single-country models include household members’ net care attitudes, net handling practices such as folding up hanging nets when not in use and the type of net user (children or adults), the type of sleeping space over which a net is used and whether households cook or store food in rooms used for sleeping [9–15]. Secondary analysis of data from 37 durability monitoring sites identified folding up hanging nets, net care attitudes and exposure to net care messages, and household size (number of members) as determinants with the largest independent effects on survival [5]. These factors are cited in the broader literature on ITN durability, including from randomized trials and qualitative studies [16–22].

The survival differences observed for the same ITN products across sites suggests that factors associated with net handling, net care attitudes and net use environment could be as predicative or more predictive of net durability than the RD score alone. Until now there has been no standardized approach to assess the combined risk to ITNs in a local setting. This has made comparisons of underlying drivers of net durability across sites and products difficult and has prevented behavioural, social, and environmental risks from being prospectively incorporated into decision-making. A single measure of risk, combining the most important determinants of ITN survival, could complement the RD score in predicting ITN performance in a sub-national setting.

Such a measure – the risk index (RI) – was previously proposed by author AK while supporting durability monitoring in the 2010s. This study presents a proof of principle of the proposed RI to quantify extrinsic durability risks and predict ITN survival using data from 44 sites across 15 countries. By so doing, it lays the foundation for investigating the RI further, to maximise its potential as a tool for vector-control decision making.

## Methods

### Data sources

This secondary analysis used data from durability monitoring studies initiated between 2015 and 2021, following the design of the United States President’s Malaria Initiative (U.S. PMI), as detailed on www.durabilitymonitoring.org. The dataset covered 44 monitoring sites across 15 countries in sub-Saharan Africa, with 14 different ITN brands assessed. Data for seven of the 44 sites were collected as part of the New Nets Project [23].

To briefly describe the primary data collection: in each site, a representative cohort of 363 (median) ITNs distributed during a mass campaign was created and monitored for up to 36 months. Surveys took place 1-6 months after distribution (when the cohort was created), and then at 12, 24 and 36 months. At each survey, the fate of cohort ITNs no longer in the household’s possession was recorded and the physical integrity of remaining cohort ITNs was assessed. These measures were combined to define the percentage of ITNs still in serviceable condition at each time point, which was used to estimate median ITN survival in months using standard graphical methods (see below). The survey questionnaire also captured durability risk factors at the individual and household levels.

For the data compiled for this analysis, four rounds of data collection occurred at 32 of the 44 sites. Data collection ended after round three (24 months) at three sites in the PMI-funded studies and for all seven New Nets Project sites. Separately, in two PMI-funded studies, the first round of data collection occurred 12 months after the campaign and only three rounds of data were collected over 36 months.

Ethical review was received for each original study prior to implementation and informed consent was attained from representatives of participating households. No ethical review was required for this secondary analysis of deidentified data.

### The Risk index

The RI was initially developed by authors AK and HK in 2017/2018 from ITN durability monitoring results from 12 sites in five countries [24]. In the present study, the original RI definition was used without modification to the original variable selection, weighting, and calculation method.

The RI was constructed as a weighted average of 12 indicators divided into three categories, representing net handling behaviours, the net use environment and net care attitudes (Table 1). The RI is an area measure, defined for each monitoring site from the mean indicator values estimated for all eligible cohort ITNs, respondents or households in a site (depending on each indicator’s unit of measure). Indicator values were taken from the baseline survey, conducted 1-6 months after the mass campaign. Refer to the supplementary information for extracts from the baseline questionnaire showing calculations for the 12 RI indicators.

**Table 1.** Indicator components of the risk index and their weights at the individual and category levels.

| Category and indicator<sup>1</sup> | Within-category weight | Sub-indicator weight | Category weight |
| --- | --- | --- | --- |
| **Net handling** | **100** |  | **45** |
| Households ever store food in sleeping room | 5 |  |  |
| Households ever cook in sleeping room | 5 |  |  |
| ITNs hanging up | 10 |  |  |
| ITNs not tied/folded when hanging | 60 |  |  |
| ITNs dried on fence/bush | 20 |  |  |
| **Net care attitudes** | **100-x** |  | **45** |
| Respondents recall message “care for your net” | 5 |  |  |
| Respondents recall message “repair your net” | 5 |  |  |
| Respondents with net care attitude score > 1.0 | 90 |  |  |
| **Net use environment** | **100** |  | **10** |
| Households with rudimentary walls (grass/mud) | 10 |  |  |
| Households cook with firewood | 30 |  |  |
| Households with rodents seen in past 6 months | 10 |  |  |
| ITNs used over a given type of sleeping place*: | 50 | 100 |  |
| Over bedframe |  | 10 |  |
| Over mattress |  | 30 |  |
| Over mat or ground |  | 60 |  |
| <sup>1</sup> All indicators except (*) are percentages ranging from 0-100 (i.e. proportions); (*) is a percentage ranging from 0 to 60. The unit of analysis is stated at the start of each indicator and is one of household, ITN or respondent (synonymous with household). |  |  |  |

Eleven of the 12 indicators are binary proportions, operationalised as percentage values ranging from 0 to 100. The RI component *ITNs used over a given type of sleeping space* was calculated as a weighted average of the composition of sleeping places over which cohort ITNs are used in the household; this summary value is then weighted at 50% when calculating the net use environment category score. The individual component values range from 0 to 60 based on the sleeping space composition. Within the net care attitudes category, the component *respondents with net care attitude score > 1.0* was based on results from a set of seven Likert scale questions with values between −2 (strongly disagree) and +2 (strongly agree). For each household, the mean value from the 7 questions was calculated and the RI component taken as the percentage of households with a mean score greater than 1.0. RI category scores were calculated as the weighted average of category indicators and the summary RI score was calculated by applying weights to the three category scores (Table 1). For the net handling and net use environment categories and their component indicators, higher values on the scale 0-100 represent greater risk of physical damage to an ITN. The components of the net care attitudes category are factors protective of ITNs and so this category is expressed as a risk by defining the score as 100 minus the weighted indicator values.

Within each group, the weights of each indicator were initially decided by AK based on plausibility, considering their importance based on current knowledge and thinking. AK then fine-tuned weights by varying them and determining the best explanatory fit on the durability outcomes from 12 monitoring sites.

### Analysis

The outcome for this analysis was median ITN survival in months. This corresponds to the time at which 50% of the cohort ITNs were no longer useable, measured from the date of ITN distribution. The outcome was interpreted graphically from empirical estimates of ITN survival in serviceable condition measured at each survey round and plotted against parametric net decay curves defined originally as part of the NetCALC tool as described by Koenker [25] and Bhatt [26]. By varying the loss function parameters, a set of decay curves was developed representing ITN median survival of one to five years, in half-year increments. Median ITN survival in months was estimated as the relative position of the last-measured data point on a horizontal line between the two adjacent theoretical median survival curves. The last round of data collection occurred 36 months after distribution in 32 sites and 24 months after distribution in 12 sites.

To describe the distribution of RI component scores, indicator values for each of the 44 sites were plotted and summarised with boxplots presenting the 25^th^, 50^th^ (median) and 75^th^ quartile values. Unweighted data were used for the descriptive analysis. Bivariable linear regression was used to assess the association between RI score and median ITN survival in months at the site level, for survival measured graphically from the last study time point. Site data were weighted to represent the number of ITNs remaining in the cohort at the last study time point. Model fit was assessed using the adjusted R^2^ value. A multivariable linear regression was subsequently performed, including the RD score for each ITN brand, to account for differences in each product’s intrinsic physical performance. As study sites are local clusters within countries, the country term was included as a random effect in the multivariable regression. Where presented, confidence intervals were calculated using the Wald method from robust standard errors. The multivariable regression also considered interactions between the RI and RD score. For ease of interpretation, results are reported as the change in median survival in months associated with a 10-point increase in the RI (and a 10-point decrease in the RD score, in the multivariable model). Regression analysis used frequency-weighted data, with the weights defined by the number of cohort ITNs remaining in the study at the final data collection time point. Analyses were performed using R v4.4.2 and StataNow v18.5 (StataCorp, College Station, Texas).#

## Results

### Risk index descriptive summary

RI scores varied from 25.1 to 83.7 (range: 58.6) across the 44 sites, with a median value of 58.9 (Figure 1). There was a relatively stable contribution of the net use environment category (mean: 5.2, standard deviation (sd): 0.9, coefficient of variation: 17%) compared to the net handling (mean: 24.5, sd: 7.5, CV: 31%) and net care attitudes (mean: 28.5, sd: 9.2, CV: 32%) categories. Decompositions of the three weighted category scores by their components are presented as supplementary information (Figure S2).

**Figure 1.**
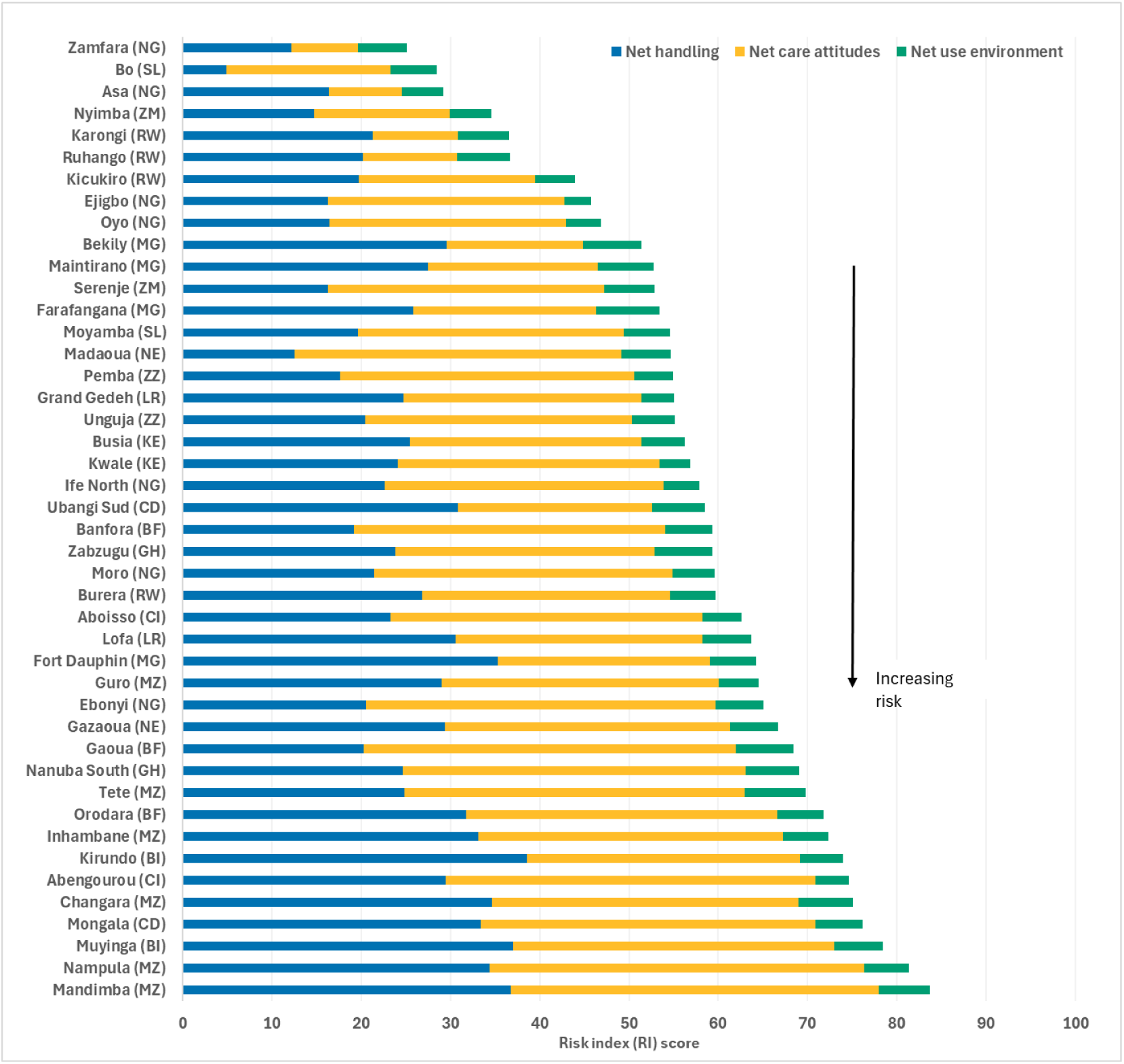
Risk index score for 44 study sites decomposed by weighted category scores using the weights in. **Table 1**. Study sites are ordered from lowest to highest RI score. RI scores are calculated from data captured during the baseline survey round conducted 1-6 months after the ITN distribution, except for Fort Dauphin, Madagascar and Serenje, Zambia where the baseline round occurred after 12 months. Standard two-letter country codes are indicated for each study site; ZZ corresponds to Zanzibar.

There was substantial site-level variation in the mean values of the 12 indicator components across the three categories (Figure 2). Ten of the twelve variables (83%) had a range exceeding 70 percentage points. There was comparatively less variation in site values for the percentage of households that ever cook in sleeping rooms (67 points), and percentage of households that use firewood for fuel (64 points). Interquartile ranges (IQRs) varied from 15 points (percentage of respondents who recall net repair message) to 43 points (percentage of household with rudimentary walls).

**Figure 2.**
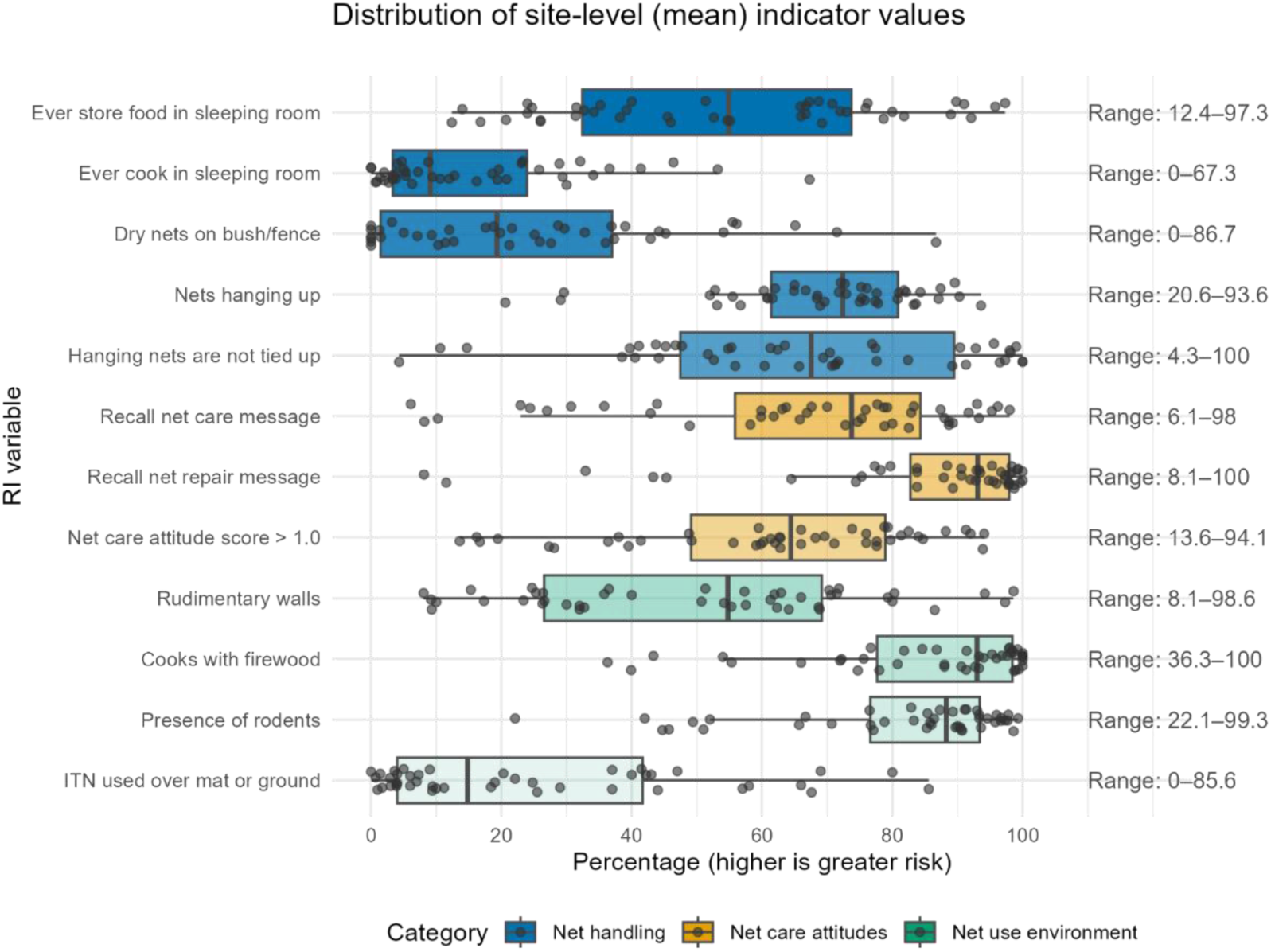
Distribution of 44 site-level (mean) values for the 12 components of the risk index score. Components are grouped by RI score category. Values for components of the net care attitude category are shown as (100 – value) reflecting their contribution to the RI score (higher value indicates greater risk). Summary box plots show 25^th^, 50^th^ (median) and 75^th^ quartile values; whiskers terminate at the values no further from the 25^th^ quartile minus 1.5 * IQR and the 75^th^ quartile plus 1.5 * IQR. For each component, point values and summary box plots have been plotted independently.

### Association of RI with median ITN survival

The association between RI and median ITN survival in months was assessed using bivariate and multivariate linear regressions (Table 2, Figure 3). In a weighted bivariate analysis, a ten-point decrease in the RI score (i.e. lower risk) was associated with an additional four months median ITN survival (95% CI: 1.7-6.3, p=0.001). The category scores were all independently associated with survival (supplementary material Figure S3). We assessed the relationship in a multivariate mixed effects model that included the ITN product RD score as a fixed effect and the study country as a random effect.

**Figure 3.**
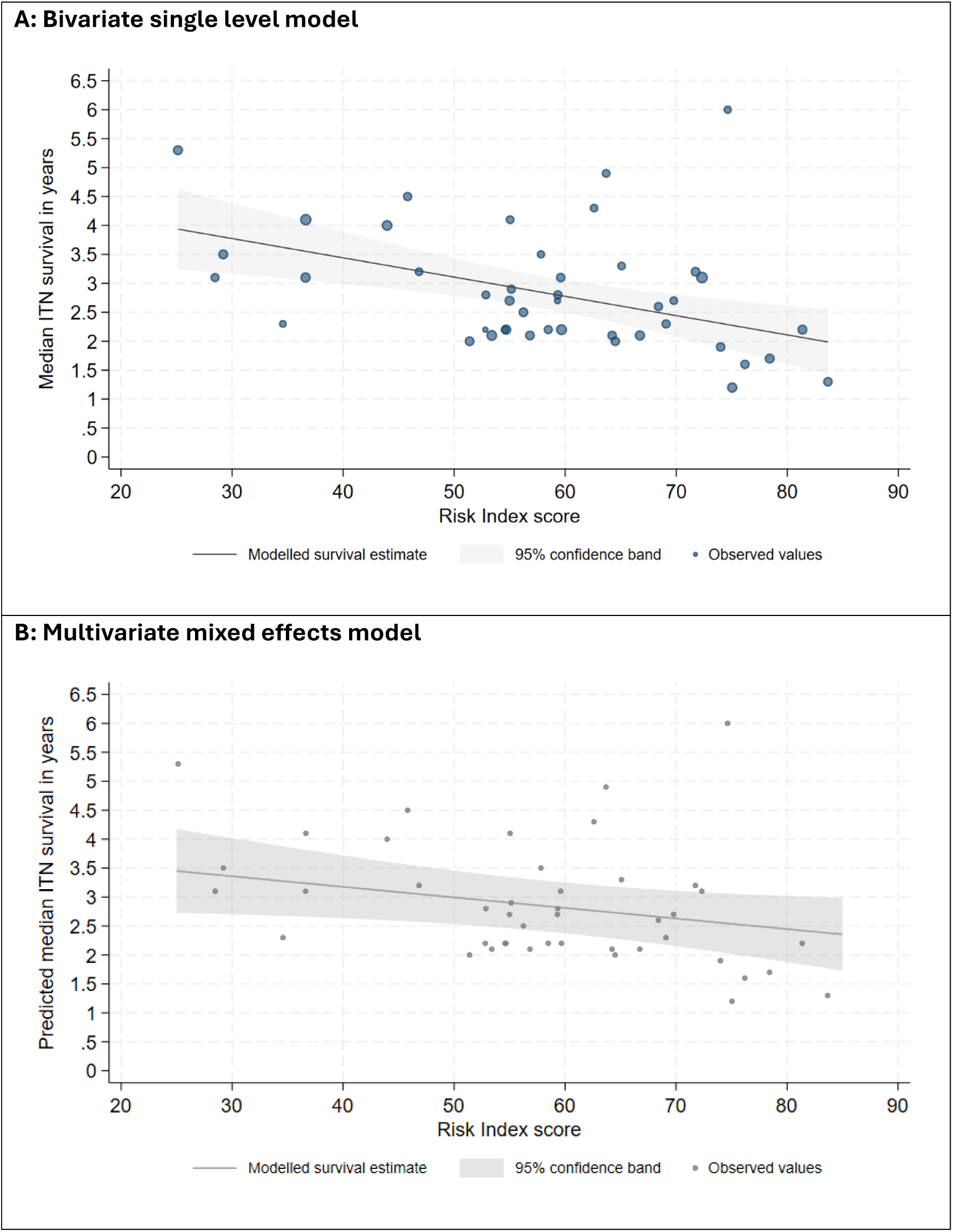
Empirical and modelled estimates of risk index score and median ITN survival. ITN median survival modelled for the range of observed RI values. **A:** Bivariate model predicted survival, estimated as the linear best fit of the risk index score on median ITN survival; data are weighted by the number of cohort ITNs active in the study at the final data collection timepoint. **B:** Multivariate model predicted survival, fitted at the mean RD score value of the observed data (55.8).

**Table 2.** Bivariate linear model and multivariate mixed-effects model for median ITN survival in months.

| Independent variable | Bivariate single level model |  |  | Multivariate mixed effects model |  |  |
| --- | --- | --- | --- | --- | --- | --- |
|  | Coefficient | s.e. | p-value | Coefficient | s.e. | p-value |
| 10-point decrease in RI | 4.00 | 1.14 | 0.001 | 2.19 | 1.05 | 0.037 |
| 10-point increase in RD | - | - | - | 3.51 | 1.10 | 0.001 |
| Constant | 57.28 | 6.85 | <0.001 | 27.32 | 10.00 | 0.006 |
| **Variance and model fit** |  |  |  |  |  |  |
| Adjusted R<sup>2</sup> | 0.207 | - | - | - | - | - |
| Variance (Constant) | - | - | - | 92.32 | - | - |
| Variance (Residual) | - | - | - | 38.91 | - | - |
| ICC | - | - | - | 0.703 | - | - |
| LR test vs linear model | - | - | - | p<0.0001 | - | - |

Bivariate model is a weighted linear model of the effect of RI on median ITN survival in months.
Multivariate model is an unweighted mixed effects model of median ITN survival in months with fixed effects of RI and RD, and county random effects.
s.e. = standard error; RI = risk index; RD = resistance to damage score [7]; ICC = intraclass correlation coefficient; LR = likelihood ratio test against a linear model without random effects

There was no evidence of an interaction between RI and RD scores (p=0.225), and no interaction term was included in the multivariate model. In this multivariate model, a ten-point decrease in the RI score corresponded to a 2 month increase in median survival (95% CI: 0.1-4.2, p=0.037); median ITN survival increased by 3.5 months for every ten-point increase in the ITN product’s RD score, independent of the RI value (95% CI: 1.3-5.7, p=0.001).

In this model, predicted median ITN survival was one year longer for the lowest observed RI value than for the lowest (Figure 3B). After accounting for RI and RD scores, differences between countries explained 70% of the remaining variance in median ITN survival.

## Discussion

This study demonstrates that the previously proposed RI metric predicts median ITN survival when applied to data from 44 sites across 15 countries in sub-Saharan Africa, representing 14 ITN brands. A ten-point decrease in the RI score corresponded to two months additional median survival. For RI scores in the range 25 to 84, this is equivalent to an additional year of survival in the site with lowest risk (Zamfara, Nigeria) compared to the highest risk (Mandimba, Mozambique). The significant association between higher RI values and reduced median ITN survival supports the predictive validity of the RI as a metric of net durability risk.

While the association between RI and median ITN survival was partially attenuated after accounting for each product’s physical durability as measured by the RD score, RI remained associated with reduced survival, supporting its relevance as an independent measure of risk to ITN lifespan. The absence of interaction between the RD score and RI is notable. This finding suggests that deploying products with higher RD scores would yield consistent survival benefits regardless of level of contextual risk. However, context can constrain the limits of survival: our results suggest that even highly durable ITNs (i.e. products with scores at the top of the current RD range) deployed in sites with high RI scores may struggle to survive up to three years (Figure 4). This programmatically important finding emphasizes the need for integrated approaches to vector control planning that address both product quality and risk context.

**Figure 4.**
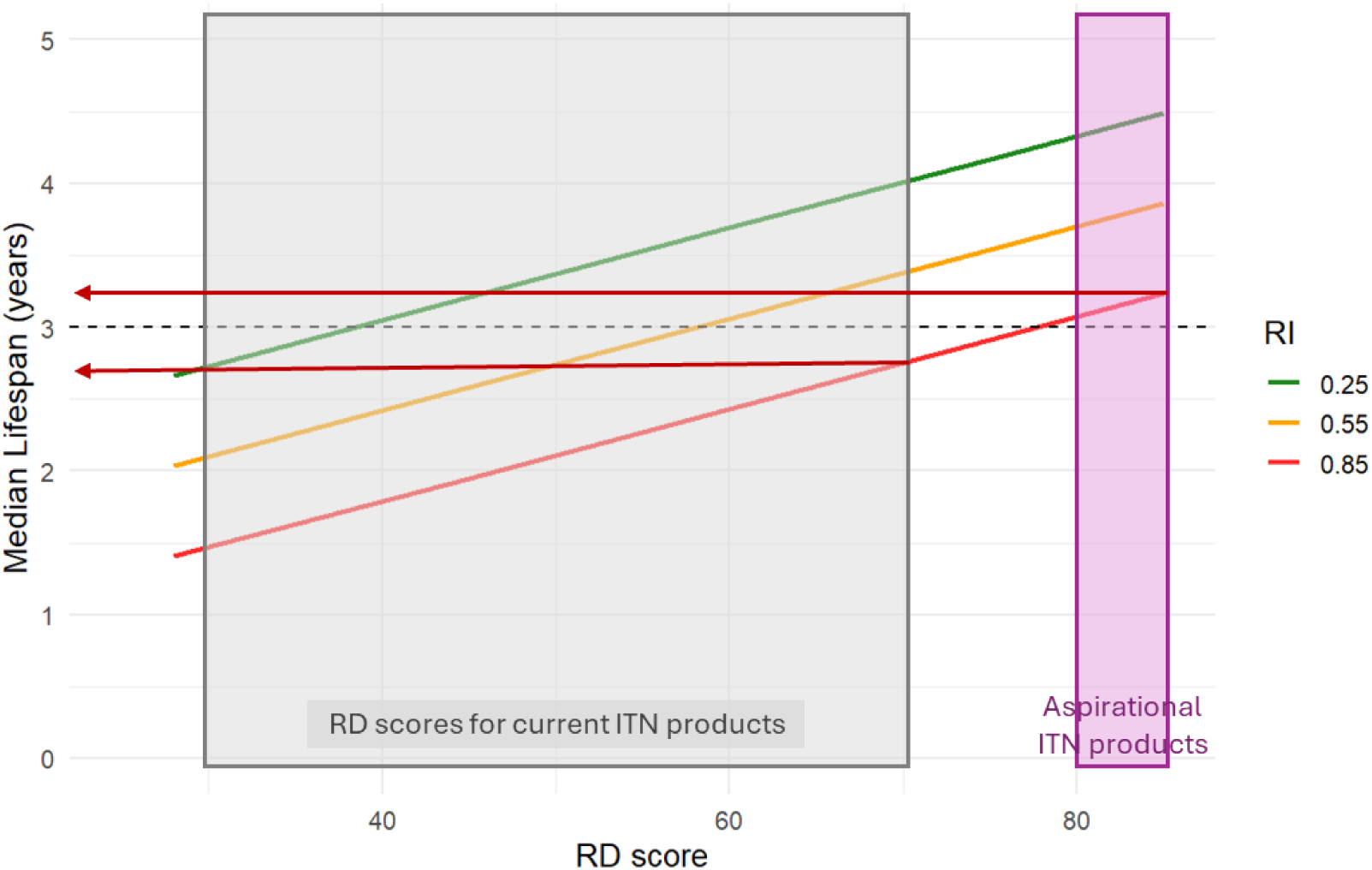
Contribution of RI and RD scores to predicted ITN median survival. Schematic representation of survival predicted by the multivariate mixed-effects model, fitted at “low” (green), “medium” (orange) and “high” (red) RI values for a range of RD scores. Currently available ITN products have RD scores in the range 35 to 70. Model predictions suggest that an ITN product at the top of the current RD range will not achieve three-year survival in settings with relatively high RI values. In such settings, three-year survival would require ITN products with “aspirational” RD scores that extend the RD score range above its current limit.

Previous studies have demonstrated substantial heterogeneity in ITN lifespan across settings, even for the same ITN product (see supplementary information). The RI approach builds on these observations by providing a quantitative framework in which to describe these differences. The current RI components reflect empirical evidence from durability monitoring studies, as outlined in the introduction. By formalizing these relationships into a composite score, the RI offers an actionable tool for programs that allows comparison of different sites.

The RI has practical applications for malaria vector control programs operating under ever tighter funding constraints. First, sub-national areas with higher RI values could receive additional consideration of vector control interventions during sub-national tailoring exercises or other programme reviews. Such areas could be prioritized for ITN products with higher physical durability (i.e. products with higher RD scores) or social and behavioural change (SBC) interventions could be designed and deployed with the objective of improving net handling and care practices among target populations.

Alternatively, analysis may indicate that switching to an alternative vector control tool is likely to be more effective. Second, by predicting differences in ITN lifespan across sub-national areas, NMPs would be better informed to make choices on appropriate ITN distribution channels and associated procurement [27], increasing the likelihood of continuous protection for at-risk populations and increasing cost-effectiveness.

Integrating data collection for the RI components into routine national monitoring activities would standardize risk assessment across sub-national areas and expand the opportunities to leverage the RI in decision making. In the absence of systematic monitoring, ad-hoc opportunities for data collection exist through integration of RI questions in national Malaria Indicator Surveys (MIS) or future implementation of Malaria Behavioral Surveys (MBS), contingent on funding.

This study has several strengths. It employs a large multi-country dataset drawing on standardized durability monitoring surveys, ensuring consistent capture and definition of both outcomes and RI components. Although it comprises 12 inputs, the RI calculation is a simple weighted average and is easy to reproduce. To separate the effect of site-level risks from intrinsic properties of different ITN products, the analysis benefits from integrating the RD score in a multivariable analysis [7].

Further scientific investigations could refine the RI and encourage uptake for decision making. First, the set of variables and the weighting scheme used to define the current RI were based on plausible associations with ITN physical survival and limited data from 12 monitoring sites. Single-country and multi-country analysis of the determinants of ITN survival are now available against which to review variable selection. Ultimately, reducing the number of variables considered and simplifying the calculation while optimizing predictive power would reduce data collection needs and lower barriers to adoption. Second, the investigation of country-level contextual variables not currently captured by the RI is required, considering the high proportion of unexplained variance at the country-level in our model. Country-level effects are likely to be proxies for the effects of unobserved ITN use patterns, seasonality or attitudinal factors. Insight into potentially relevant factors may come from an examination of MIS and MBS data collected contemporaneously to the durability monitoring studies included in this analysis. Third, exploring how the RI scores and predictive power vary over different spatial and temporal scales will be important to match the metric to appropriate geographic units. This will also determine the optimal frequency and spatial distribution of data collection required to generate representative data at administrative levels relevant for vector control decision-making.

This study has some limitations. Considering the primary data sources, most RI components rely solely on responses from household members and as such are susceptible to recall bias and social desirability bias. Considering the estimation of median ITN survival, it is reported that some study sites experienced high levels of cohort ITNs with unknown outcomes (i.e. respondents were unsure what had happened to an ITN between survey visits). These ITNs were excluded from calculations of ITN survival. High loss to follow-up may bias survival results ITNs with unknown outcomes are systematically different from ITNs with known outcomes; the direction and scale of any difference is unknown.

## Conclusion

The previously proposed RI is a standardized metric for quantifying the combined impact of net handling behaviours, net care attitudes and the net use environment on ITN lifespan. The RI permits meaningful comparisons of the risks to ITN survival across settings and is predictive of ITN survival, independent of ITN product. Combined with the RD score, the RI could strengthen subnational tailoring of ITN strategies by responding by fitting ITN products to relative levels of risk. Future refinements of the RI will seek to simplify the metric to support uptake by NMPs and their partners.

## Data Availability

Data and results from the original durability monitoring studies are available for download from https://www.durabilitymonitoring.org/. Summary data and analysis code for this secondary analysis is available from the corresponding author upon reasonable request.

## Acknowledgments

The authors would like to acknowledge the contribution of the large number of study partner organisations and individuals across 15 countries involved in the primary data collection for durability monitoring.

## Author contributions

AK, SP and FM designed the study protocol. AK originated the risk index metric. SP conducted secondary data analysis. SP and JAT drafted the manuscript. All authors critically reviewed the manuscript. All authors read and approved the final manuscript.

## Ethics approval and consent to participate

Not applicable.

## Consent for publication

All authors have read and approved the final version of the manuscript and consent to its publication.

## Supplementary information

**Figure S1.**
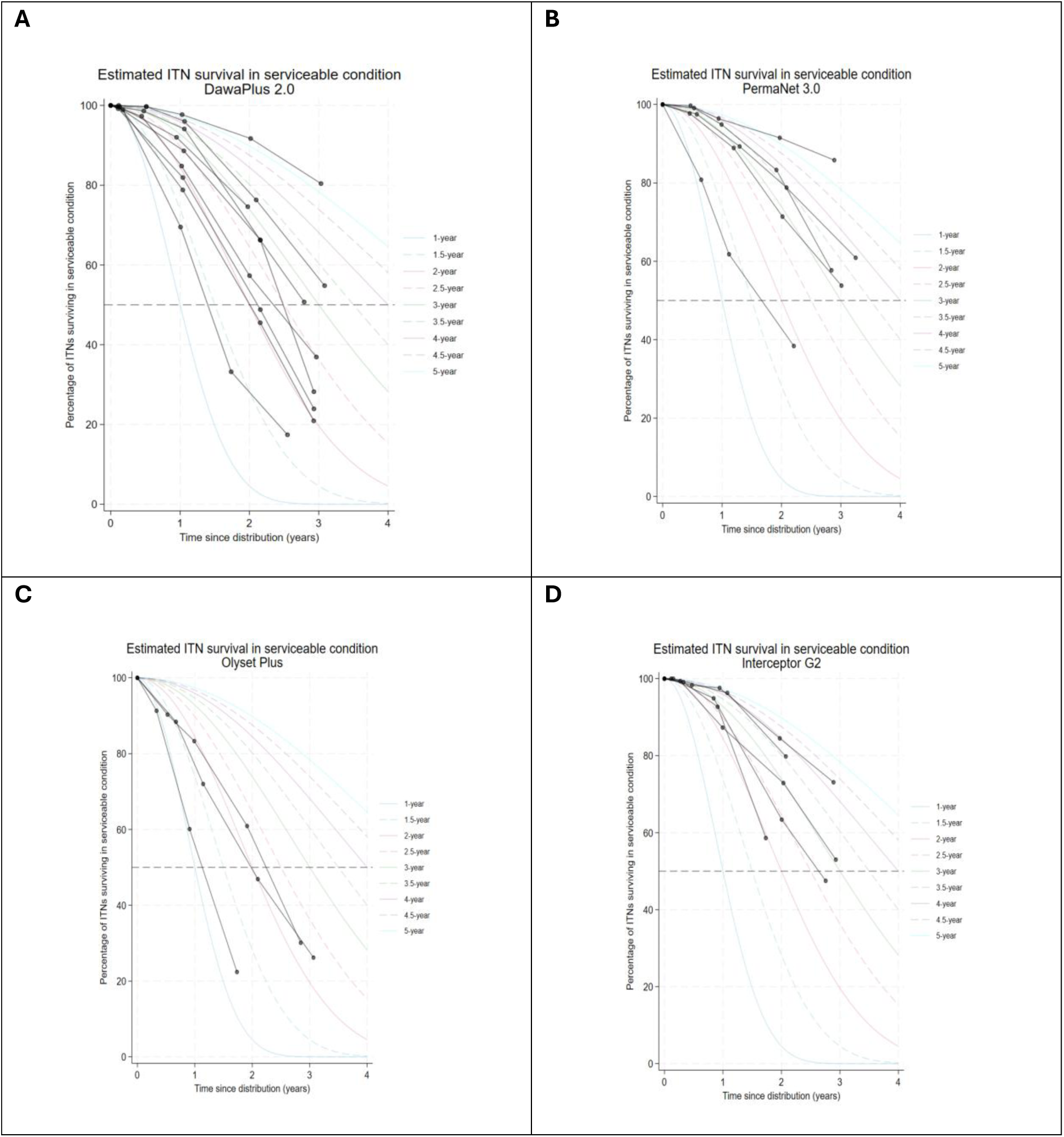
Heterogeneity in measures of ITN survival within and across countries that cannot be explained by ITN products’ intrinsic physical properties alone. Figure presents summary ITN durability monitoring survival results for four products monitored in at least three study sites. **A**: Data for DawaPlus ® 2.0 ITNs at nine sites in Democratic Republic of the Congo (DRC), Ghana, Kenya, Madagascar and Nigeria, with ITN median survival in the range 1.6 (DRC) to 5.3 years (Nigeria); **B**: PermaNet ® 3.0 ITNs at five sites in Burkina Faso, Burundi, Cote d’Ivoire, Rwanda and Sierra Leone, with ITN median survival in the range 1.9 to 6.0 years median lifespan; **C**: Olyset ® Plus ITNs at three sites in Mozambique, Sierra Leone and Zambia, with ITN median survival in the range 1.2 to 2.3 years; **D**: Interceptor ® G2 ITNs at five sites in Burkina Faso, Cote d’Ivoire, Mozambique, Nigeria and Rwanda, with ITN median survival in the range 2.0 to 4.3 years.

**Figure S2.**
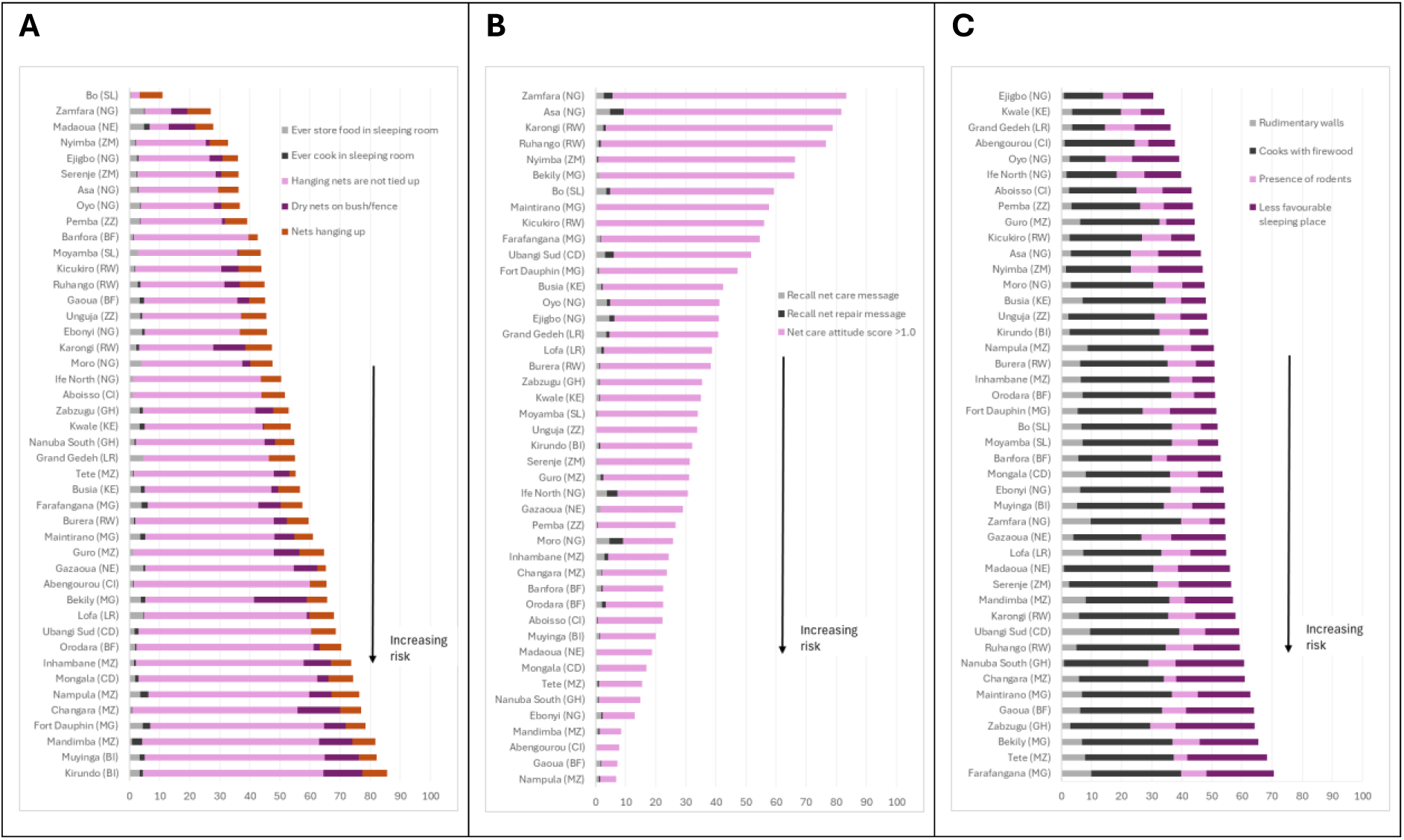
Risk index category scores for 44 study sites decomposed by weighted components using the weights in. **Table 1. A:** Net handling score; **B:** Net care attitude score (inverse), the value used in the RI calculation is 100 minus the score shown, such that lower net care attitude scores reflect greater risk; **C:** Net use environment score. Study sites are ordered from lowest to highest risk in each panel independently. Standard two-letter country codes are indicated for each study site; ZZ corresponds to Zanzibar.

**Figure S3.**
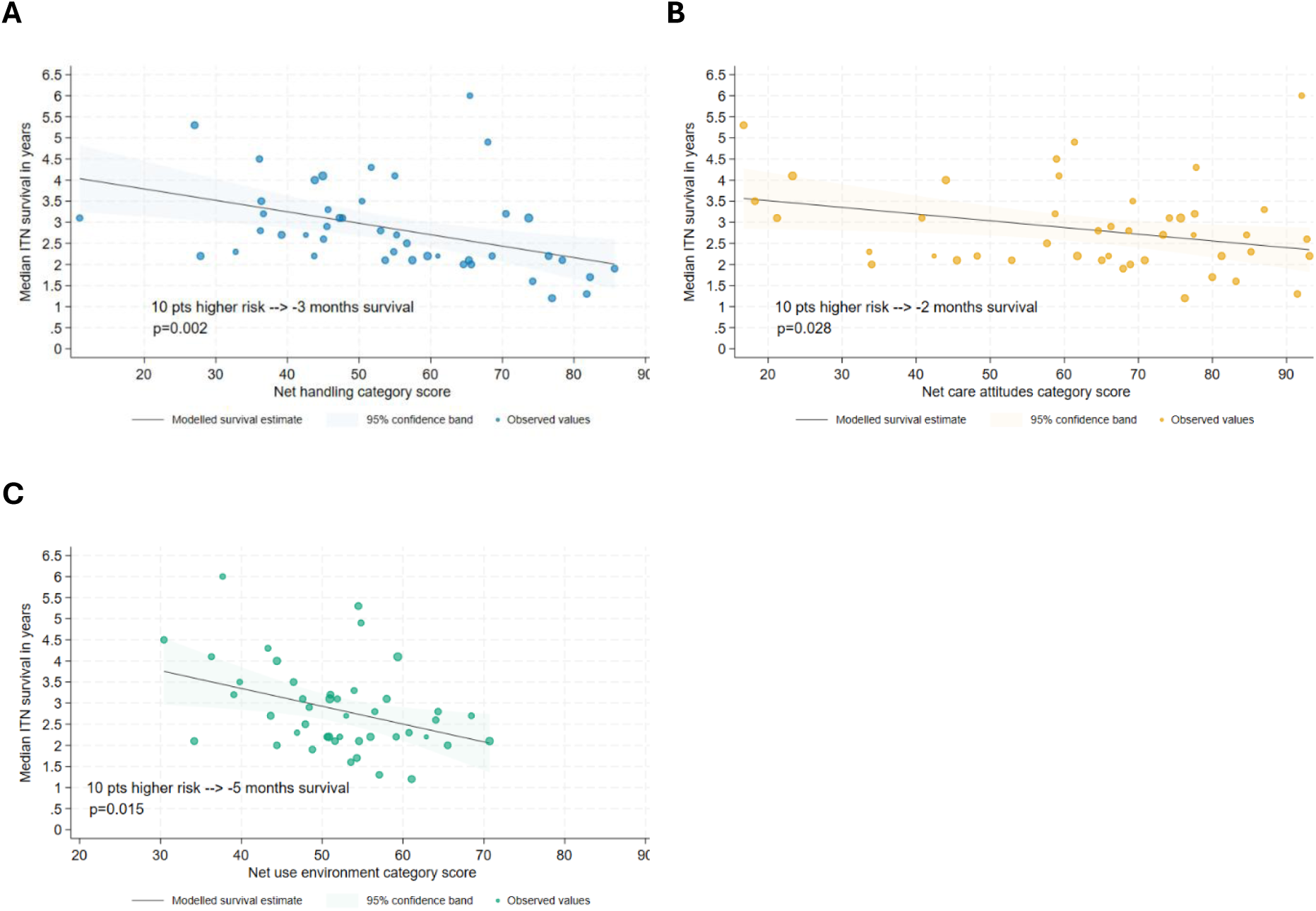
Empirical and modelled estimates of the three risk index category scores and median ITN survival. **A.** Net handling category score; **B.** Net care attitudes category score; **C.** Net use environment category score. Modelled survival is the linear best fit of the risk index category score on median ITN survival; data are weighted by the number of cohort ITNs active in the study at the final data collection timepoint. Survival is modelled for the range of observed category values.

## Definition of indicators that comprise the risk index

The 12 indicators that comprise the risk index are presented below in bold. Each indicator name is followed by relevant questions from the household survey questionnaire, guidance on how data are manipulated (where relevant) and numerator and denominator definitions.

**Net handling: Percentage of households that report ever storing food in rooms used for sleeping.**

|  |  |  |  |
| --- | --- | --- | --- |
|  | Do you ever store food in a room that is also used for sleeping? | Yes | 1 |
|  |  | No | 0 |
|  |  | Don't know | 9 |

*Numerator:* Number of households responding Yes

*Denominator:* Number of households with a non-missing response (including don’t know responses)

**Net handling: Percentage of households that report ever cooking in rooms used for sleeping.**

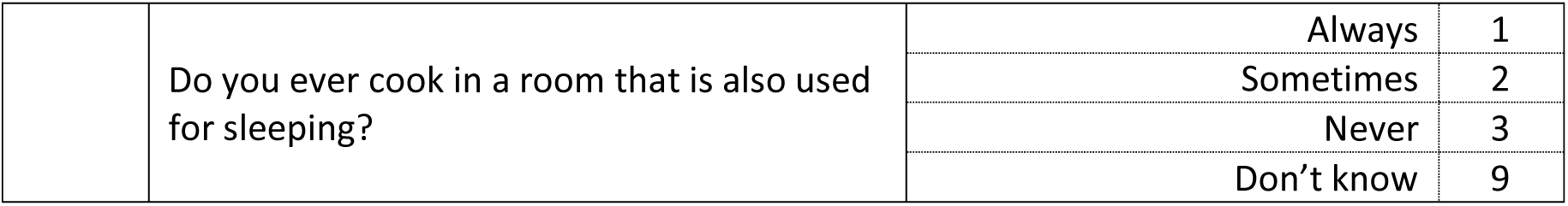

|  |  |  |  |
| --- | --- | --- | --- |
|  | Do you ever cook in a room that is also used for sleeping? | Always | 1 |
|  |  | Sometimes | 2 |
|  |  | Never | 3 |
|  |  | Don't know | 9 |

*Numerator:* Number of households responding Always or Sometimes

*Denominator:* Number of households with a non-missing response (including don’t know responses)

**Net handling: Percentage of ITNs that are hanging up**

|  |  |  |  |
| --- | --- | --- | --- |
|  | Where was the net found? | Hanging loose over sleeping place | 1 |
|  |  | Hanging and folded up or tied | 2 |
|  |  | Not hanging but not stored | 3 |
|  |  | Stored away unpacked | 4 |
|  |  | Stored away still in package | 5 |

*Numerator:* Number of ITNs with response option 1 or 2

*Denominator:* Number of ITNs with a non-missing response

**Net handling: Percentage of ITNs that are not tied or folded up when hanging**

*Numerator:* Number of ITNs with response option 1

*Denominator:* Number of ITNs with response option 1 or 2

**Net handling: Percentage of ITNs that were dried on a fence or bush after they were washed**

|  |  |  |  |
| --- | --- | --- | --- |
| A | Has this net ever been washed? | Yes | 1 |
|  |  | No | 0 |
|  |  | Don't know | 9 |
| B | **Among washed nets:**<br>For the last wash, what soap was used? | Soap bar | 1 |
|  |  | Detergent (OMO etc) | 2 |
|  |  | Bleach | 3 |
|  |  | Mix | 4 |
|  |  | None | 5 |
| C | **Among washed nets:**<br>Where was the net dried? | Outside on the ground | 1 |
|  |  | Outside on line | 2 |
|  |  | Outside bush or fence | 3 |
|  |  | Inside | 4 |

*Numerator:* Number of ITNs reportedly dried “outside bush or fence” (Option 3 to Question C)

*Denominator:* Number of ITNs that have ever been washed (Yes to Question A)

**Net care attitudes: Percentage of respondents who recall the message “care for your net”**

|  |  |  |  |
| --- | --- | --- | --- |
| A | In the last 6 months, did you receive any information on use and care and repair of your mosquito nets from any source? | Yes | 1 |
|  |  | No | 0 |
|  |  | Don’t know | 9 |
| B | What was the content of the messages you heard/saw?<br>**>> multiple answers possible** | Use your net | 1 |
|  |  | Care for your net | 2 |
|  |  | Hang up your net | 3 |
|  |  | Sleep under your net every night | 4 |
|  |  | Nets prevent malaria | 5 |
|  |  | Repair your net | 6 |
|  |  | Other | 8 |
|  |  | Specify: |  |

*Numerator:* Number of households citing “Care for your net” or similar response captured by “Other”

*Denominator:* Number of households with non-missing responses for Question A

**Net care attitudes: Percentage of respondents who recall the message “repair your net”**

*Numerator:* Number of households citing “Repair your net” or similar response captured by “Other”

*Denominator:* Number of households with non-missing responses for Question A

**Net care attitudes: Percentage of respondents with a net care attitude score > 1.0**

|  |  |  |  |  |  |
| --- | --- | --- | --- | --- | --- |
| I am going to read a series of statements to you and I would like you to tell me how much you agree with them. For each statement, please tell me if you strongly agree, somewhat agree, somewhat disagree, or strongly disagree with it. |  |  |  |  |  |
|  |  | Strongly agree | Somewhat agree | Somewhat disagree | Strongly disagree |
| LS1 | Mosquito nets are valuable | 1 | 2 | 3 | 4 |
| LS2 | There are actions I can take to make my net last long | 1 | 2 | 3 | 4 |
| LS3 | It is not possible to repair holes in nets (Reverse coded statement) | 1 | 2 | 3 | 4 |
| LS4 | A repaired net can still be effective against mosquitoes | 1 | 2 | 3 | 4 |
| LS5 | Other people in this community fix holes in their mosquito nets | 1 | 2 | 3 | 4 |
| LS6 | I do not have time to repair a hole in my net (Reverse coded statement) | 1 | 2 | 3 | 4 |
| LS7 | I can help protect my family from malaria by taking care of my net | 1 | 2 | 3 | 4 |
| LS8 | I am confident I can repair holes immediately | 1 | 2 | 3 | 4 |

Strongly agree responses are scored as +2 Somewhat agree responses are scored as +1

Somewhat disagree responses are scored as −1 Strongly disagree responses are scored as −2

For cases with non-missing responses to LS1, LS2, LS3, LS4, LS6, LS7, LS8: The average response score is calculated as:

1/7 x (LS1 + LS2 + (−1 x LS3) + LS4 + (−1 x LS6) + LS7 + LS8)

*Numerator:* Number of households with an average score > 1.0

*Denominator:* Number of households with a non-missing score

*Note:* LS5 is included in the questionnaire but excluded from the calculation

**Net use environment: Percentage of households with rudimentary walls (grass/mud)**

|  |  |  |  |
| --- | --- | --- | --- |
|  | What is the main material of the walls? | Grass | 1 |
|  |  | Mud | 2 |
|  |  | Plastered | 3 |
|  |  | Brick/Concrete | 4 |

*Numerator:* Number of households citing “Grass” or “Mud” (or other rudimentary material as relevant for each site)

*Denominator:* Number of households with non-missing responses

**Net use environment: Percentage of households that cook with firewood**

|  |  |  |  |
| --- | --- | --- | --- |
|  | What is the main energy source for cooking? | Firewood | 1 |
|  |  | Charcoal | 2 |
|  |  | Kerosene | 3 |
|  |  | Gas | 4 |
|  |  | Electricity | 5 |

*Numerator:* Number of households citing “Firewood”

*Denominator:* Number of households with non-missing responses

**Net use environment: Percentage of households that report seeing rodents in the past 6 months**

|  |  |  |  |
| --- | --- | --- | --- |
|  | In the last 6 months, have you seen any rats or mice in your house or their traces (faeces or damage)? | Yes | 1 |
|  |  | No | 0 |
|  |  | Don't know | 9 |

*Numerator:* Number of households citing “Yes”

*Denominator:* Number of households with non-missing responses (including don’t know responses)

**Net use environment: Percentage of ITNs that are used over a given type of sleeping space**

|  |  |  |  |
| --- | --- | --- | --- |
| Q88 | What type of sleeping place has this net been used for mostly? |  |  |
|  |  | Bed frame (finished) | 1 |
|  |  | Bed frame (sticks) | 2 |
|  |  | Foam mattress | 3 |
|  |  | Reed mat | 4 |
|  |  | Grass | 5 |
|  |  | Ground | 6 |
|  |  | Never used | 7 |

Bed frame response options (1 and 2) are coded to “Bedframe” Foam mattress response (3) is coded to “Mattress”

Other responses (4, 5 and 6) are coded to “Mat or ground”

*Numerator:* Number of ITNs with a given type of sleeping space

*Denominator:* Number of ITNs that have been used (i.e. are not unused) with a non-missing response

## References

1. The changing impact of malaria control in Africa 2000-2025 – MAP [Internet]. Malaria Atlas Project; [cited 2026 Mar 17]. https://malariaatlas.org/project-resources/impact-of-control-2000-2025/. Accessed 17 Mar 2026

2. World Health Organization. World malaria report 2025. Geneva: World Health Organization; 2025.

3. Ekelru J. Global Fund’s Eighth Replenishment: A Funding Chasm | GFO Aidspan [Internet]. Aidspan. [cited 2026 Mar 17]. https://aidspan.org/Blog/view/32580/global_fund_s_eighth_replenishment_a_funding_c hasm. Accessed 17 Mar 2026

4. Bertozzi-Villa A, Bever CA, Koenker H, Weiss DJ, Vargas-Ruiz C, Nandi AK, et al. Maps and metrics of insecticide-treated net access, use, and nets-per-capita in Africa from 2000-2020. Nat Commun. 2021;12:3589. 10.1038/s41467-021-23707-7

5. Poyer S, Worges M, Sternberg E. Determinants of ITN durability in sub-Saharan Africa: secondary analysis of data from 37 durability monitoring sites [Internet]. The Alliance for Malaria Prevention; 2025 [cited 2026 Mar 17]. https://allianceformalariaprevention.com/wp-content/uploads/2025/11/Durability_determinants-_ITN_survival_102025_EN.pdf. Accessed 17 Mar 2026

6. Kilian A, Obi E, Mansiangi P, Abílio AP, Haji KA, Guillemois E, et al. Correlation of textile ‘resistance to damage’ scores with actual physical survival of long-lasting insecticidal nets in the field. Malaria Journal. 2021;20:29. 10.1186/s12936-020-03570-5

7. Mechan F, Poyer S, Tangena J-AA, Worges M, Sternberg E, Kilian A, et al. Refining the resistance-to-damage (RD) score to predict operational Insecticide-Treated Net lifespan and identify paths to innovation [Internet]. bioRxiv; 2025 [cited 2026 Mar 17]. p. 2025.03.02.641027. 10.1101/2025.03.02.641027

8. Tools | LLIN Durability Monitoring [Internet]. [cited 2026 Apr 17]. https://www.durabilitymonitoring.org/?page_id=10. Accessed 17 Apr 2026

9. Sudoi RK, Esch K, Yamba F, Iyikirenga L, Youssef C, Nallo P, et al. Physical and insecticidal durability of PermaNet® 3.0 and Olyset® plus piperonyl butoxide-synergist insecticide-treated nets in Sierra Leone: results of durability monitoring from 2020 to 2023. Malaria Journal. 2025;24:230. 10.1186/s12936-025-05421-7

10. Raharinjatovo J, Dabiré RK, Esch K, Soma DD, Hien A, Camara T, et al. Physical and insecticidal durability of Interceptor®, Interceptor® G2, and PermaNet® 3.0 insecticide-treated nets in Burkina Faso: results of durability monitoring in three sites from 2019 to 2022. Malar J. 2024;23:173. 10.1186/s12936-024-04989-w

11. Obi E, Okoh F, Blaufuss S, Olapeju B, Akilah J, Okoko OO, et al. Monitoring the physical and insecticidal durability of the long-lasting insecticidal net DawaPlus® 2.0 in three States in Nigeria. Malaria Journal. 2020;19:124. 10.1186/s12936-020-03194-9

12. Haji KA, Khatib BO, Obi E, Dimoso K, Koenker H, Babalola S, et al. Monitoring the durability of the long-lasting insecticidal nets Olyset® and PermaNet® 2.0 in similar use environments in Zanzibar. Malaria Journal. 2020;19:187. 10.1186/s12936-020-03258-w

13. Mansiangi P, Umesumbu S, Etewa I, Zandibeni J, Bafwa N, Blaufuss S, et al. Comparing the durability of the long-lasting insecticidal nets DawaPlus® 2.0 and DuraNet© in northwest Democratic Republic of Congo. Malaria Journal. 2020;19:189. 10.1186/s12936-020-03262-0

14. Abílio AP, Obi E, Koenker H, Babalola S, Saifodine A, Zulliger R, et al. Monitoring the durability of the long-lasting insecticidal nets MAGNet and Royal Sentry in three ecological zones of Mozambique. Malaria Journal. 2020;19:209. 10.1186/s12936-020-03282-w

15. Hiruy HN, Irish SR, Abdelmenan S, Wuletaw Y, Zewde A, Woyessa A, et al. Durability of long-lasting insecticidal nets (LLINs) in Ethiopia. Malar J. 2023;22:109. 10.1186/s12936-023-04540-3

16. Morgan J, Abílio AP, Pondja M do R, Marrenjo D, Luciano J, Fernandes G, et al. Physical Durability of Two Types of Long-Lasting Insecticidal Nets (LLINs) Three Years After a Mass LLIN Distribution Campaign in Mozambique, 2008–2011. The American Journal of Tropical Medicine and Hygiene. The American Society of Tropical Medicine and Hygiene; 2015;92:286–93. 10.4269/ajtmh.14-0023

17. Hunter GC, Scandurra L, Acosta A, Koenker H, Obi E, Weber R. “We are supposed to take care of it”: a qualitative examination of care and repair behaviour of long-lasting, insecticide-treated nets in Nasarawa State, Nigeria. Malar J. 2014;13:320. 10.1186/1475-2875-13-320

18. Scandurra L, Acosta A, Koenker H, Kibuuka DM, Harvey S. “It is about how the net looks”: a qualitative study of perceptions and practices related to mosquito net care and repair in two districts in eastern Uganda. Malar J. 2014;13:504. 10.1186/1475-2875-13-504

19. Massue DJ, Moore SJ, Mageni ZD, Moore JD, Bradley J, Pigeon O, et al. Durability of Olyset campaign nets distributed between 2009 and 2011 in eight districts of Tanzania. Malar J. 2016;15:176. 10.1186/s12936-016-1225-6

20. Lorenz LM, Bradley J, Yukich J, Massue DJ, Mboma ZM, Pigeon O, et al. Comparative functional survival and equivalent annual cost of 3 long-lasting insecticidal net (LLIN) products in Tanzania: A randomised trial with 3-year follow up. PLOS Medicine. Public Library of Science; 2020;17:e1003248. 10.1371/journal.pmed.1003248

21. Solomon T, Loha E, Deressa W, Balkew M, Gari T, Overgaard HJ, et al. Bed nets used to protect against malaria do not last long in a semi-arid area of Ethiopia: a cohort study. Malar J. 2018;17:239. 10.1186/s12936-018-2391-5

22. Tomass Z, Alemayehu B, Balkew M, Leja D. Knowledge, attitudes and practice of communities of Wolaita, Southern Ethiopia about long-lasting insecticidal nets and evaluation of net fabric integrity and insecticidal activity. Parasites Vectors. 2016;9:224. 10.1186/s13071-016-1494-5

23. New Nets Project: Evidence of effectiveness and cost-effectiveness of dual-AI ITNs from the observational pilot studies—final report [Internet]. [cited 2026 Mar 31]. https://www.path.org/our-impact/resources/new-nets-project-evidence-of-effectiveness-and-cost-effectiveness-of-dual-ai-itns-from-the-observational-pilot-studiesfinal-report/. Accessed 31 Mar 2026

24. Kilian A, Koenker H. Can durability of Long-lasting Insecticidal Nets be predicted from a ‘Risk Index’ composed of selected variables on net handling and use environment? 2018.

25. Koenker HM, Yukich JO, Mkindi A, Mandike R, Brown N, Kilian A, et al. Analysing and recommending options for maintaining universal coverage with long-lasting insecticidal nets: the case of Tanzania in 2011. Malar J. 2013;12:150. 10.1186/1475-2875-12-150

26. Bhatt S, Weiss DJ, Mappin B, Dalrymple U, Cameron E, Bisanzio D, et al. Coverage and system efficiencies of insecticide-treated nets in Africa from 2000 to 2017. Kyobutungi C, editor. eLife. 2015;4:e09672. 10.7554/eLife.09672

27. Alliance for Malaria Prevention (AMP). Guidance on channel selection for distribution of insecticide-treated nets [Internet]. 2025 Aug. https://allianceformalariaprevention.com/wp-content/uploads/2025/10/Channel_selection_guide_092025_EN.pdf. Accessed 17 Apr 2026

